# A score-based risk model for predicting severe COVID-19 infection as a key component of lockdown exit strategy

**DOI:** 10.1101/2020.05.20.20108571

**Authors:** Noa Dagan, Noam Barda, Dan Riesel, Itamar Grotto, Siegal Sadetzki, Ran Balicer

## Abstract

**Background:** As many countries consider and employ various lockdown exit strategies, health authorities seek tools to provide differential targeted advice for social distancing based on personal risk for severe COVID-19. However, striking a balance between a scientifically precise multivariable risk prediction model, and a model which can easily be used by the general public, remains a challenge. A list of risk criteria, as defined by the CDC for example, provides a simple solution, but may be too inclusive by classifying a substantial portion of the population at high risk. Score-based risk classification tools may provide a good compromise between accuracy and simplicity.

**Objective:** To create a score-based risk classification tool for severe COVID-19.

**Methods:** The outcome was defined as a composite of being labeled severe during hospitalization or dying due to COVID-19. The risk classification tool was developed using retrospective data from all COVID-19 patients that were diagnosed until April 1^st^, 2020 in a large healthcare organization (“training set”). The developed tool combines 10 risk factors using simple summation, and defines three risk levels according to the patient’s age and number of accumulated risk points – basic risk, high risk and very-high risk (the last two levels are also considered together as the elevated risk group). The tool’s performance in accurately identifying individuals at risk was evaluated using a “temporal test set” of COVID-19 patients diagnosed between April 2^nd^ and April 22^nd^, 2020, later than those used for model development. The tool’s performance was also compared to that of the CDC’s criteria. The healthcare organization’s general population was used to evaluate the proportion of patients that would be classified to each of the model’s risk levels and as elevated risk by the CDC criteria.

**Results:** A total of 2,421, 2,624 and 4,631,168 individuals were included in the training, test, and general population cohorts, respectively. The outcome rate in the training and test sets was 5%. Overall, 18% of the general population would be classified at elevated risk by the model, with a resulting sensitivity of 92%, compared to 35% that would be defined as elevated risk by the CDC criteria, with a resulting sensitivity of 96%. Within the model’s elevated risk groups, the high and very-high risk groups comprised 15% and 3% of the general population, with an incidence rate (PPV) of 15% and 33%, respectively.

**Discussion:** A simple to communicate score-based risk classification tool classifies at elevated risk about half of the population that is considered to have an elevated risk by the CDC risk criteria, with only a 4% reduction in sensitivity. The model’s ability to further divide the elevated risk population into two markedly different subgroups allows providing more refined recommendations to the general public and limiting the restrictions of social distancing to a smaller and more manageable subset of the population. This model was adopted by the Israeli ministry of health as its risk classification tool for COVID-19 lab tests prioritization and for targeting its instructions on risk management during the lockdown exit strategy.

## Introduction

During the early phases of the COVID-19 pandemic, many countries applied extended and highly inclusive confinement measures, including mass lockdown^1^, to halt disease dissemination. As many countries consider and employ various lockdown exit strategies, health authorities seek tools to provide differential advice for social distancing based on, among other factors, personal risk for severe COVID-19.

As more data accumulated on the outcome of COVID-19 patients, a growing list of risk factors for severe disease and mortality has been suggested. The Centers for Disease Control and Prevention (CDC) put forth a list of criteria to define people at high risk for severe COVID-19 disease. This list includes those 65 years of age and older, those who live in nursing homes or long-term care facilities, and those who meet at least one of a long list of clinical criteria^2^. However, while being straightforward and simple to use, such an extensive list of singular criteria tends to be too inclusive, classifying a large proportion of the population as high-risk individuals.

Multivariable prediction models can help define a more refined and accurate classification of risk^3^. However, most of these models cannot be easily communicated to the public to allow self-identification of populations at risk.

A third option, that fits conceptually on the complexity spectrum between single-dimension risk criteria and multivariable prediction models, are simplified score-based risk models. These models, which have long been used in medicine (e.g. Wells criteria^4^, CHADS2-Vasc^5^), strike a good balance between simplicity and accuracy and therefore are highly applicable for daily routine decisions.

The need for a score-based model to stratify risk for severe COVID-19 illness, one that could both be easily communicated to lay people and readily used by medical professionals, was deemed critical by the Israeli Ministry of Health for the country’s exit strategy. This paper describes the process by which this risk model was developed and evaluated. The evaluation was performed using a temporal validation strategy, with data that was collected after the model’s training period. The paper also compares the score-based model performance in accurately identifying high-risk individuals with those of the CDC risk criteria.

## Methods

### Setting and Data Sources

This is a retrospective cohort study based on the data of Clalit Health Services (CHS) and the Israeli Ministry of Health (MOH).

CHS is Israel’s largest integrated payer-provider healthcare organization. Health insurance in Israel is universal, provided to all residents by one of four such integrated organizations. CHS covers over half of the Israeli population (4.6 million members), providing them with primary and specialty care, laboratory testing, imaging studies and hospital services. The CHS central data warehouse includes over 20 years of electronic health records and claims-based data and is a unique resource for planning and research.

During the COVID-19 pandemic, the Israeli MOH established centralized nation-wide data collection and reporting services, recording the performance and results of all COVID-19 lab tests, as well as COVID-19 related hospital admissions. The report also includes a daily status for each patient, ranked as mild, moderate or severe (at the treating physician’s discretion), and documentation of every mortality event. These data were also shared with the health organizations, where they were cross-linked with over 20 years of existing patient records. This combined repository was used in this study.

### Study Population and outcome definition

The dataset for this study comprised of all CHS members as of February 1^st^, 2020. Nested in this group was the training dataset which included all patients diagnosed with COVID-19 at or before April 1^st^, 2020.

A second, temporally separate dataset (‘validation set’) included all patients that were diagnosed between April 2^nd^, 2020 and April 22^nd^, 2020 (three weeks prior to the data extraction date, which was May 13^th^, 2020), thus allowing at least three weeks of follow-up. No patients were lost to follow-up.

The outcome of interest was a composite of COVID-19 related death or COVID-19-related admission labeled as “severe” at any point after diagnosis. Secondary analyses assessed the mortality outcome alone.

### Model Development

To select covariates for the model, we identified the key features associated with increased mortality among lab-confirmed COVID-19 cases in the largest population-based cohort study published to date^6^. Other covariates were selected from recently published lists of risk factor for severe COVID-19 illness^7^, from risk factors that were identified by a multivariate prediction model that was previously developed and implemented in the CHS ^3^ and from known risk factors for other respiratory diseases^8^. To facilitate public sharing of the model, an important consideration when choosing covariates was their explainabilty to the non-medical population and their expected availability to healthcare providers. This consideration led to the inclusion of health conditions and health behaviors, and the exclusion of information that the general population may not know such as lab results.

The final list of variables chosen to be included in the model were cardiovascular disease or congestive heart failure, diabetes mellitus, overweight (BMI ≥ 30), active or recent malignancy, immunosuppression, chronic obstructive pulmonary disease or over 10 smoking pack-years, chronic hepatic, renal or neurological disease and hospital admissions in the last 3 years (with the exception of those for normal delivery). The selected list of covariates was then extracted for all PCR lab-confirmed COVID-19 patients. Covariates were extracted no later than a month prior to diagnosis, to prevent data leaks resulting from the suspected or confirmed COVID-19 status. A full technical description of the covariates and outcomes, and the way they were used to construct the risk categories, is included in Supplemental Table 1.

Next, we determined the criteria for the risk classification. Considerations regarding ease of use of the model by the lay public led to the decision to assign a single point for each risk factor (previous admissions were allotted 1 point per admission), stratified by age. Age was found to be the main driver of risk of severe disease among those infected^7^, and age groups were thus addressed as an interaction variable in this model, sub-classified by the total number of risk points accumulated. Patients who were missing data in a variable were simply not assigned points for that variable.

Outcome incidence rates were determined for each 10-year age group, and these were clustered into broader age categories of similar risk. Then, a heatmap of the outcome rate in the intersection between each age category and points count was plotted and used to create three risk levels – basic risk, high risk and very-high risk.

The very-high risk group was to receive relatively strict home isolation recommendations (unless low rates of disease dissemination are present), and was therefore set to include a relatively small proportion of the population, with only the highest rates of severe illness - no less than 25%. The basic risk group was to receive the minimal set of restrictions given by the government as general advice, and accordingly was to include the vast majority of the population, with low rates of severe illness - no more than 5%. The population subgroups with outcomes rates higher than 5%, but still substantially lower than the very-high risk group, were defined as ‘high risk’.

### Model Validation

The model’s performance was assessed on a “temporal” validation dataset that included only patients diagnosed later than the patients in the training set. The model was assessed for sensitivity, specificity, positive predictive value (PPV) and negative predictive value (NPV) in each stratum of risk, and also for all patients at elevated risk (i.e. the very-high risk and high risk groups combined). A 95% confidence interval (CI) for each measure was derived using the exact binomial distribution. Performance was further compared to that of the CDC’s high-risk criteria.

As the lab confirmed COVID-19 infected population is likely not representative of the entire population, we also evaluated the proportion of the general population assigned to each risk strata. This was done by applying the model’s classification, as well as the CDC criteria, to the entire CHS population as of February 1, 2020.

### Ethical approval

This study was approved by Clalit Health Services’ institutional review board (0052-20-COM).

### Analysis

All analysis was performed using R version 3.5.2.

## Results

The entire CHS population comprised 4,631,168 individuals as of February 1^st^, 2020. From these, a total of 110,310 members (2.4%) were tested for COVID-19 between February 1^st^, and April 22^rd^, 2020, with a total of 5,045 (4.6%) positive cases (a population flow chart is presented in Figure 1).

**Figure 1.**
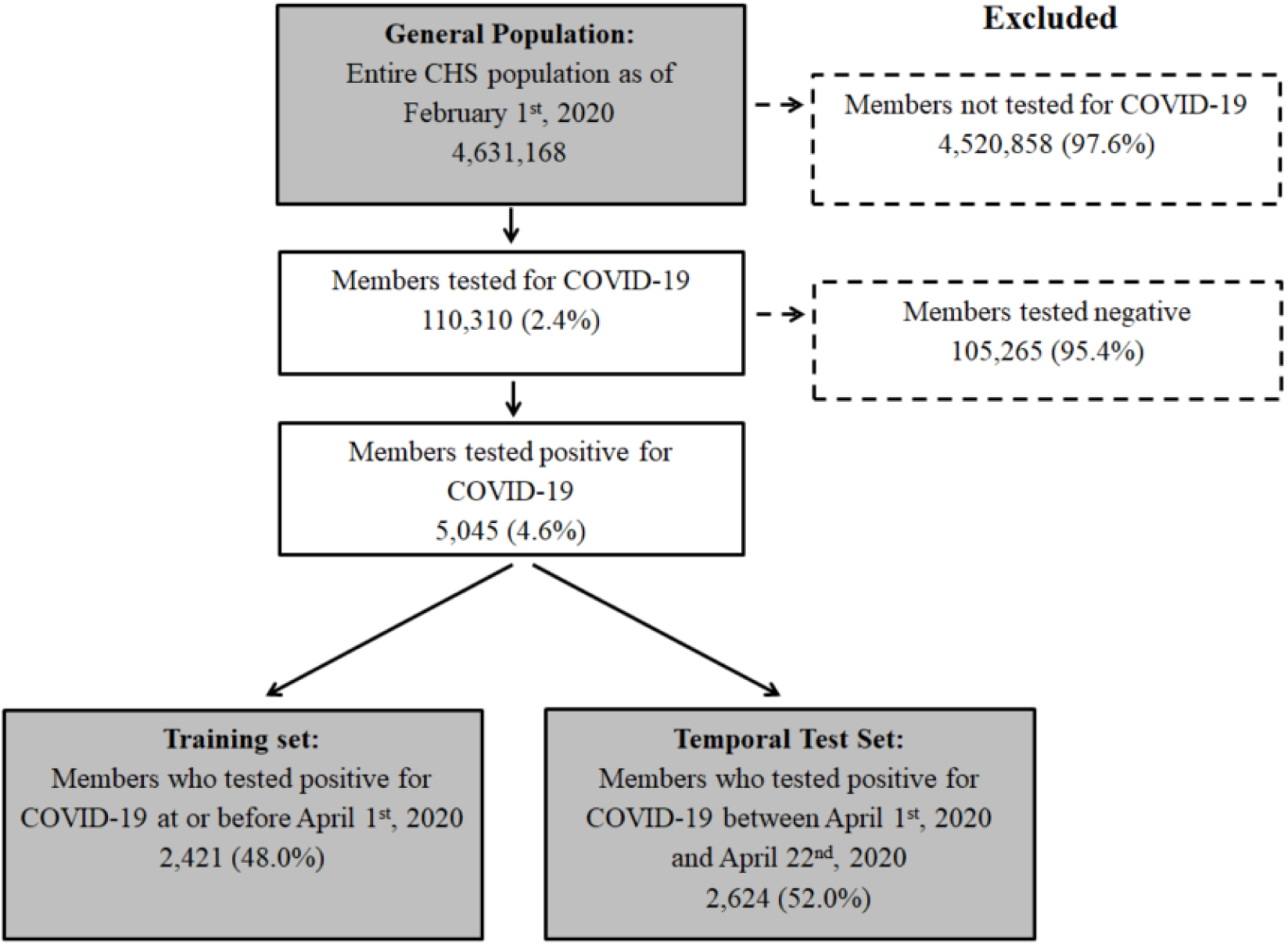
– Population flow chart.

The model’s training set included 2,421 laboratory confirmed COVID-19 patients. Over the follow-up period, 122 (5.0%) patients experienced the composite outcome and 75 (3.1%) the secondary death outcome (Table 1).

**Table 1.** – Population characteristics table.

| Variable | General Population dataset | Training Set (lab-confirmed up to April 1<sup>st</sup>) | Temporal Test Set (lab-confirmed between April 1<sup>st</sup> and May 8<sup>th</sup>) | Missing in Train/Test Sets |
| --- | --- | --- | --- | --- |
| **Overall** | 4,631,168 | 2,421 | 2,624 |  |
| **Main outcome – composite of death and severe state, n (%)** |  |  |  | 0% |
| No | -- | 2,299 (95.0) | 2,489 (94.9) |  |
| Yes | -- | 122 (5.0) | 135 (5.1) |  |
| **Secondary outcome – death, n (%)** |  |  |  | 0% |
| No | -- | 2,346 (96.9) | 2,554 (97.3) |  |
| Yes | -- | 75 (3.1) | 70 (2.7) |  |
| **Age, n (%)** |  |  |  | 0% |
| 0-9 | 938,864 (20.3) | 110 (4.5) | 213 (8.1) |  |
| 10-19 | 708,741 (15.3) | 199 (8.2) | 338 (12.9) |  |
| 20-29 | 593,328 (12.8) | 547 (22.6) | 510 (19.4) |  |
| 30-39 | 642,565 (13.9) | 376 (15.5) | 358 (13.6) |  |
| 40-49 | 517,709 (11.2) | 316 (13.1) | 304 (11.6) |  |
| 50-59 | 385,460 (8.3) | 281 (11.6) | 272 (10.4) |  |
| 60-69 | 397,934 (8.6) | 316 (13.1) | 254 (9.7) |  |
| 70-79 | 268,428 (5.8) | 166 (6.9) | 161 (6.1) |  |
| 80-89 | 142,609 (3.1) | 78 (3.2) | 142 (5.4) |  |
| 90+ | 35,530 (0.8) | 32 (1.3) | 72 (2.7) |  |
| **CVD / CHF, n (%)** |  |  |  | 0% |
| No | 4,305,025 (93.0) | 2,179 (90.0) | 2,344 (89.3) |  |
| Yes | 326,143 (7.0) | 242 (10.0) | 280 (10.7) |  |
| **Diabetes Mellitus, n (%)** |  |  |  | 0% |
| No | 4,225,611 (91.2) | 2,145 (88.6) | 2,297 (87.5) |  |
| Yes | 405,557 (8.8) | 276 (11.4) | 327 (12.5) |  |
| **Obesity, n (%)** |  |  |  | 4.9% |
| No | 3,468,027 (83.5) | 1,827 (78.8) | 1,903 (76.7) |  |
| Yes | 685,112 (16.5) | 491 (21.2) | 578 (23.3) |  |
| **Active/Recent Malignancy, n (%)** |  |  |  | 0% |
| No | 4,531,378 (97.8) | 2,342 (96.7) | 2,559 (97.5) |  |
| Yes | 99,790 (2.2) | 79 (3.3) | 65 (2.5) |  |
| **Chronic Renal Disease, n (%)** |  |  |  | 0% |
| No | 4,365,351 (94.3) | 2,237 (92.4) | 2,418 (92.1) |  |
| Yes | 265,817 (5.7) | 184 (7.6) | 206 (7.9) |  |
| **Chronic Hepatic Disease, n (%)** |  |  |  | 0% |
| No | 4,566,137 (98.6) | 2,369 (97.9) | 2,570 (97.9) |  |
| Yes | 65,031 (1.4) | 52 (2.1) | 54 (2.1) |  |
| **Chronic Neurological Disease, n (%)** |  |  |  | 0% |
| No | 4,438,872 (95.8) | 2,286 (94.4) | 2,408 (91.8) |  |
| Yes | 192,296 (4.2) | 135 (5.6) | 216 (8.2) |  |
| **Immunosuppression, n (%)** |  |  |  | 0% |
| No | 4,538,015 (98.0) | 2,365 (97.7) | 2,560 (97.6) |  |
| Yes | 93,153 (2.0) | 56 (2.3) | 64 (2.4) |  |
| **COPD / Smoking 10+ Pack Years, n (%)** |  |  |  | 0% |
| No | 3,997,267 (86.3) | 2,135 (88.2) | 2,352 (89.6) |  |
| Yes | 633,901 (13.7) | 286 (11.8) | 272 (10.4) |  |
| **3Y Hospital admission Count,** |  |  |  | 0% |

| n (%) |  |  |  |
| --- | --- | --- | --- |
| 0 | 3,885,063 (83.9) | 2,010 (83.0) | 2,063 (78.6) |
| 1 | 489,671 (10.6) | 241 (10.0) | 312 (11.9) |
| 2 | 139,946 (3.0) | 84 (3.5) | 114 (4.3) |
| 3 | 54,374 (1.2) | 35 (1.4) | 50 (1.9) |
| 4+ | 62,114 (1.3) | 51 (2.1) | 85 (3.2) |

Abbreviations: CVD, Cardiovascular Disease; CHF, Congestive Heart Failure; COPD, Chronic Obstructive Pulmonary Disease

After considering the outcome rate in each 10-year age group (Figure 2a), the age groups were combined into broader age categories of similar risk – ages 0-29, 30-49, 50-69 and 70 years or higher. Based on a heatmap that considered the interaction between the chosen age categories and the number of points (Figure 2b) with the predefined thresholds for the risk levels detailed above, the final cutoff points for each risk level were defined (Figure 2c).

**Figure 2.**
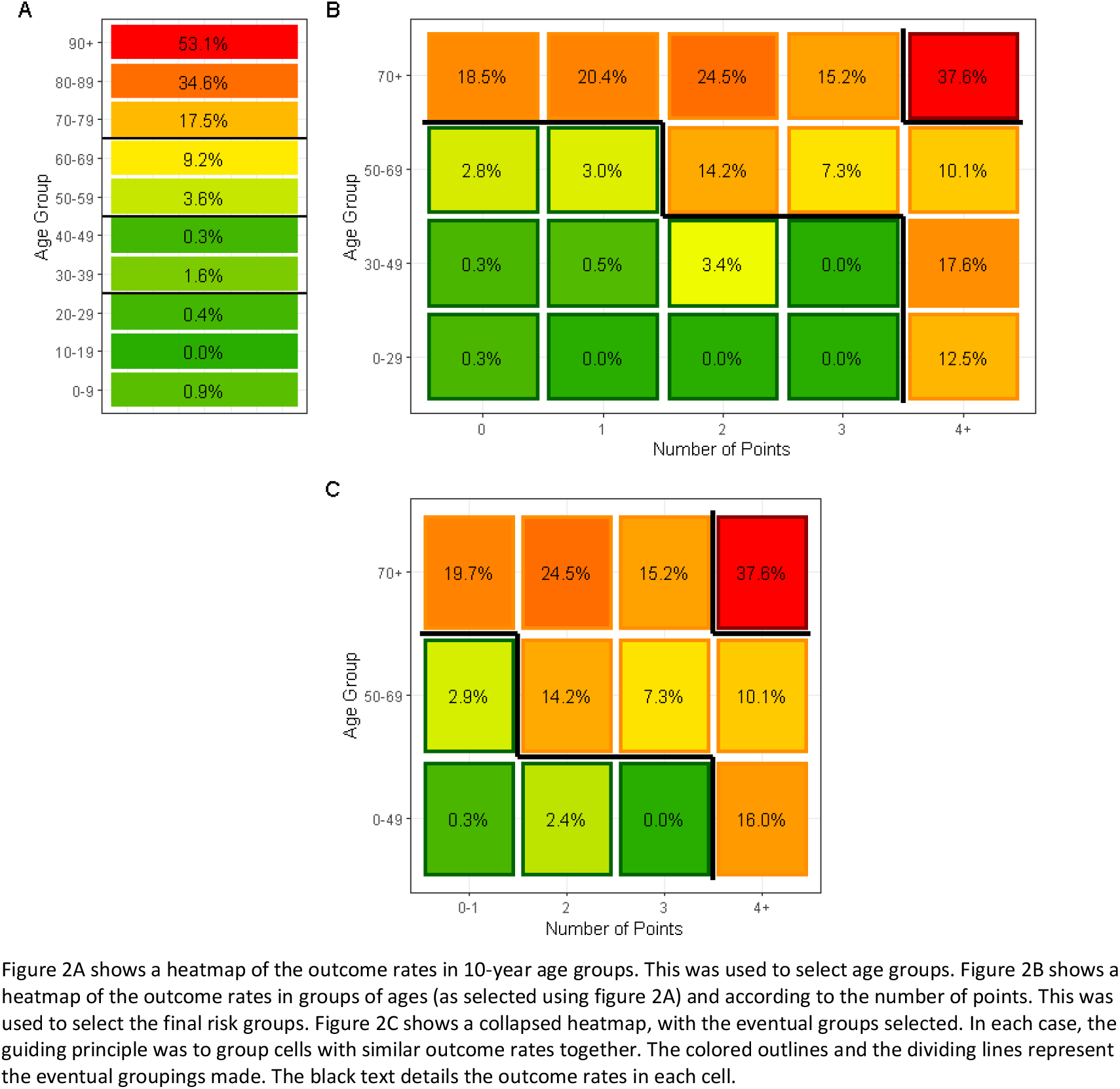
– Risk for severe course of disease or death by age groups and point count.

Figure 3 presents a summary of the final model; Patients of any age with four or more risk points, patients aged 50-69 years with 2 or more risk points and patients over the age of 70 years with 0-3 risk points were classified as the high-risk group. Patients over the age of 70 years with 4 or more risk points were classified as the very-high risk group. All other patients were classified as the basic risk group.

**Figure 3.**
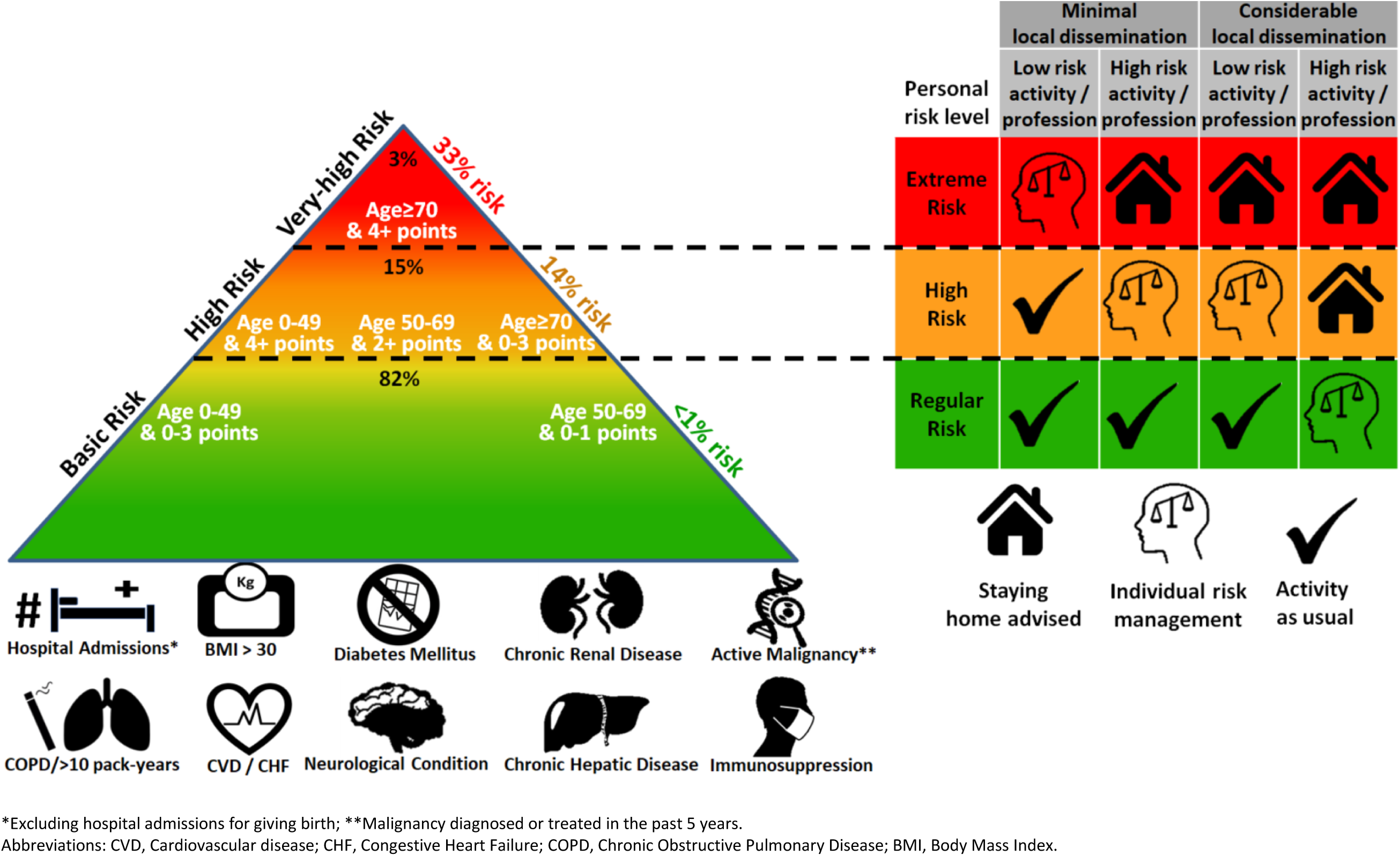
– Summary of the score-based model and policy recommendations for individual risk management.

According to this model, a total of 35.4% of the entire CHS population (50.4% over the age of 20) will be classified as elevated risk by the CDC criteria, compared to 18.0% (27.6% over the age of 20) using the score-based model (considering the very-high and high risk groups combined) (Table 2). Among the 2,624 lab-confirmed COVID-19 patients in the temporal test set (Table 1), a total of 96.3% (95% CI: 91.6-98.8%) and 91.9% (95% CI: 85.9-95.9%) of the 122 patients with the composite outcome were correctly identified as elevated risk (sensitivity) by the CDC criteria and score-based model, respectively (Table 2). In addition, of those who would have been classified as elevated risk by the CDC criteria and by the model, a total of 13.1% (95% CI: 11.0-15.3%) and 19.6% (95% CI: 16.5-22.9%) would have suffered severe COVID-19 or death, respectively. Both models identified all death cases as elevated risk.

**Table 2.** – Model Performance.

| All age groups |  |  |  |  |  |  |  |  |  |  |
| --- | --- | --- | --- | --- | --- | --- | --- | --- | --- | --- |
|  |  |  | Main outcome – Severe COVID-19 cases and death |  |  |  | Secondary outcome – COVID-19 deaths |  |  |  |
|  | Percent included in the group of all CHS members (95% CI) | Percent included in the group among COVID-19 lab confirmed patients (95% CI) | Sensitivity % (95% CI) | PPV % (95% CI) | Specificity % (95% CI) | NPV % (95% CI) | Sensitivity % (95% CI) | PPV % (95% CI) | Specificity % (95% CI) | NPV % (95% CI) |
| CDC risk criteria | 35.4 (35.4-35.4) | 37.9 (36-39.8) | 96.3 (91.6-98.8) | 13.1 (11.0-15.3) | 65.3 (63.4-67.2) | 99.7 (99.3-99.9) | 100.0 (94.9-100) | 7.0 (5.5-8.8) | 63.8 (61.9-65.7) | 100.0 (99.8-100.0) |
| Elevate risk* | 18.0 (17.9-18.0) | 24.2 (22.5-25.8) | 91.9 (85.9-95.9) | 19.6 (16.5-22.9) | 79.5 (77.9-81.1) | 99.4 (99-99.7) | 100.0 (94.9-100) | 11.0 (8.7-13.7) | 77.9 (76.3-79.5) | 100.0 (99.8-100) |
| Very-high Risk | 3.0 (2.9-3.0) | 6.8 (5.9-7.8) | 43.0 (34.5-51.8) | 32.6 (25.8-40) | 95.2 (94.3-96) | 96.9 (96.1-97.5) | 54.3 (41.9-66.3) | 21.3 (15.6-28.1) | 94.5 (93.6-95.4) | 98.7 (98.2-99.1) |
| High Risk | 15.0 (15.0-15.0) | 17.4 (15.9-18.9) | 48.9 (40.2-57.6) | 14.5 (11.4-18) | 84.3 (82.8-85.7) | 96.8 (96-97.5) | 45.7 (33.7-58.1) | 7.0 (4.8-9.8) | 83.4 (81.9-84.8) | 98.2 (97.6-98.8) |
| Basic Risk | 82.0 (82.0-82.1) | 75.8 (74.2-77.5) | 8.1 (4.1-14.1) | 0.6 (0.3-1) | 20.5 (18.9-22.1) | 80.4 (77.1-83.5) | 0.0 (0.0-5.1) | 0.0 (0.0-0.2) | 22.1 (20.5-23.7) | 89.0 (86.3-91.3) |
| Ages 20 years and older |  |  |  |  |  |  |  |  |  |  |
|  |  |  | Main outcome – Severe COVID-19 cases and death |  |  |  | Secondary outcome – COVID-19 deaths |  |  |  |
|  | Percent included in the group of all CHS members (95% CI) | Percent included in the group among COVID-19 lab confirmed patients (95% CI) | Sensitivity % (95% CI) | PPV % (95% CI) | Specificity % (95% CI) | NPV % (95% CI) | Sensitivity % (95% CI) | PPV % (95% CI) | Specificity % (95% CI) | NPV % (95% CI) |
| CDC risk criteria | 50.4 (50.3-50.5) | 46.0 (43.8-48.1) | 96.3 (91.6-98.8) | 13.6 (11.5-16) | 57.5 (55.3-59.7) | 99.6 (99-99.9) | 100.0 (94.9-100.0) | 7.3 (5.8-9.2) | 55.9 (53.7-58.1) | 100.0 (99.7-100.0) |
| Elevate risk* | 27.6 (27.5-27.6) | 30.5 (28.5-32.5) | 91.9 (85.9-95.9) | 19.6 (16.6-22.9) | 73.8 (71.8-75.7) | 99.2 (98.6-99.6) | 100.0 (94.9-100.0) | 11.1 (8.7-13.8) | 71.9 (69.9-73.9) | 100.0 (99.7-100.0) |
| Very-high Risk | 4.6 (4.6-4.6) | 8.6 (7.4-9.9) | 43.0 (34.5-51.8) | 32.6 (25.8-40.0) | 93.8 (92.6-94.8) | 95.9 (94.9-96.8) | 54.3 (41.9-66.3) | 21.3 (15.6-28.1) | 93.0 (91.8-94.1) | 98.3 (97.6-98.8) |
| High Risk | 23.0 (23.0-23.1) | 21.9 (20.1-23.7) | 48.9 (40.2-57.6) | 14.5 (11.4-18.1) | 80.0 (78.1-81.7) | 95.7 (94.6-96.7) | 45.7 (33.7-58.1) | 7.0 (4.9-9.8) | 78.9 (77.1-80.7) | 97.7 (96.8-98.3) |
| Basic Risk | 72.4 (72.4-72.5) | 69.5 (67.5-71.5) | 8.1 (4.1-14.1) | 0.8 (0.4-1.4) | 26.2 (24.3-28.2) | 80.4 (77.1-83.4) | 0.0 (0.0-5.1) | 0.0 (0.0-0.3) | 28.1 (26.1-30.1) | 88.9 (86.2-91.3) |

\* Very-high & High risk groups combined.
Abbreviations: PPV, Positive Predictive Value; NPV, Negative Predictive Value; CDC, Centers for Disease Control and prevention; CI, Confidence Interval

The score-based model further stratifies the elevated risk group into very-high risk and high-risk groups. The model designates 3.0% of the CHS population in the very-high risk group, achieving a sensitivity of 43.0% (95% CI: 34.5-51.8%) and a PPV of 32.6% (95% CI: 25.8-40.0%). The high-risk group included 15.0% of the CHS population, with a sensitivity of 48.9% (95% CI: 40.2-57.6%) and PPV of 14.5% (95% CI: 11.4-18.0%). The basic risk group as defined by the score-based model, which included 82.0% of the total population had an outcome rate of 0.6% (95% CI: 0.3-1.0%) for the composite outcome and 0% (95% CI: 0-0.2%) for the death outcome.

## Discussion

In this study we presented a relatively simple score-based model to classify the risk of patients for severe COVID-19 illness into three levels. The simplicity of the model facilitates its use by both medical professionals and the lay public for decision-making regarding COVID-19 prevention and treatment. The model performs well, correctly identifying 92% of patients who will experience a severe COVID-19 infection or death as having an elevated risk, while only classifying 18% of the total population as such. This is in contrast to the CDC’s list of risk criteria, which classify nearly two times as many patients (35%) at elevated risk, for a small 4% gain in sensitivity (96%).

The model further classifies the elevated risk group to very-high risk and high risk groups, which comprise 3% and 15% of the population, respectively. These groups are significantly different from one another – while the first is relatively small, it captures 43% of all individuals that will be severely ill. On average, 33% of the patients defined as very-high risk will have severe illness if infected, and 21% will die. The high risk group is considerably larger, and captures 49% of all severely ill patients, and these patients have a 15% risk on average to be severely ill, and 7% risk of mortality. This division of the elevated risk population into two markedly different subgroups, allows health authorities to provide more refined recommendations, and to limit more restrictive social distancing recommendations to a smaller and more manageable subset of the population (Figure 3).

In order for a tool to provide useful personal risk classification in the outpatient setting, it must be highly selective in its inclusion criteria. Specifically, the need articulated by the policy makers in Israel was for a tool that would, if possible, identify over 90% of severe cases among those infected, while labeling 20% or less of the population as individuals with an elevated risk. The target sensitivity of 90% was set in order to provide one order of magnitude reduction in the number of severe cases, and thus an order of magnitude reduced strain on ICU beds (assuming that these identified individuals will be adequately risk-avoidant during the lockdown exit strategy).

This target cannot be achieved by considering every risk factor for severe illness as a sufficient criterion for defining a person as high risk. As shown in this study, this is indeed the case when the CDC risk factor list is employed to define high-risk individuals – it defines as such 35% of the entire CHS member population and over 50% of all adults. As a single risk factor in some age groups entails a negligible absolute risk^6^, a practical risk classification method has to address the personal absolute risk driven by multiple concomitant risk factors in the context of a person’s age, as done by the suggested risk-points based approach.

An earlier version of this score-based tool was used in Israel’s national COVID-19 strategy since April 14^th^, 2020. Early on, the national guidelines for COVID-19 testing were based on this risk classification. Additionally, as part of the national lockdown exit strategy, this score-based risk model allowed for more refined, stratified government advice on social distancing. This was done with an emphasis that the model cannot replace patient-specific medical judgment, as it may not “pick up” on less common conditions that still pose an obvious risk to some patients. Specifically, it was used to define which teachers and students would continue staying home even after frontal education was resumed.

Currently, more detailed and dynamic guidelines are being considered by the Israeli MOH. These guidelines refine the social distancing recommendation to be based on three axes (as depicted in Figure 3). The first axis considers the individual’s risk of severe disease if infected, which is provided by the score-based model. This risk should be considered in light of the temporospatial level of disease dissemination (the second axis); In Israel, the MOH designed a method to define “red” areas based on the level and trend of positive test results. In this way, individuals can easily consider the disease activity in their area and integrate this factor into their personal risk management. Finally, individual risk management should also take into account a person’s pattern of activities and their associated likelihood of contact with infected individuals (the third axis). High risk professions such as school-teachers and high risk activities such as providing medical care should be considered in a different manner. Conveying a complicated message such as individual risk management based on three axes to the general public is challenging but necessary. Figure 3 was constructed as a visual aid to educate the general public on how to perform this individual risk management themselves, taking location, day and specific activity into account.

The methodology used in this study was derived from the predefined aim of creating a user-friendly and intuitive tool that could be communicated to the lay public for personal use. For this purpose, a “one-point” per risk factor approach was adopted. Of all these risk factors, one factor, the number or recent hospital admission, stands out in its separate scoring. The choice to assign each hospital admission a separate point was made for three reasons – first, this variable was found to be highly indicative of high risk when considering thousands of candidate variables^3^; second, its value reflects the severity of the other chronic conditions that were uniformly assigned a single point, thus compensating to some extent for not accounting for the variability of these conditions’ risk level; and third, it sometimes stands for rarer conditions that were not included as specific risk factors in the model.

Other risk classification methods have been suggested^9,10^, most of which based on prediction models that cannot be manually calculated. Xie et al^11^ and Gong et al^12^ developed prediction models for severe COVID-19 and translated them into a graphical nomograms. Both models are designed to assess risk of inpatients, and accordingly use measures that are only available in a hospital setting. Also, the nomogram’s complexity is more suitable for medical staff than for lay people. Unlike these models, the point-based model suggested in this study is meant to help decide on risk mitigation strategies prior to contracting SARS-CoV-2. For these purposes, the model must be based solely on background characteristics and on a cohort that includes both COVID-19 outpatients and inpatients. The availability of data regarding background characteristics and outcomes for all COVID-19 patients in the CHS makes the development of such a tool possible.

The presented score-based risk model was developed to provide a useful compromise between effective risk classification and simplicity of use – this model achieved a sensitivity of over 90%, while flagging fewer than 20% of the population as elevated risk. This risk model is proving helpful to the Israeli health authorities in providing recommendations on lab testing allocation and stratified guidance on social distancing and self-isolation. Owing to the model’s concise and user friendly format, it can easily be shared and validated across countries and healthcare providers, enabling more nuanced population health maintenance policies, point-of-care decisions, and individual empowerment in individual risk management. Further validation in multiple countries and settings is needed to ascertain the model’s performance in different populations.

## Data Availability

Access to the data used for this study can be made available upon request, subject to an internal review by RB to ensure that participant privacy is protected, and subject to completion of a data sharing agreement, approval from the institutional review board of Clalit Health Services and institutional guidelines and in accordance with the current data sharing guidelines of Clalit Health Services and Israeli law. Pending the aforementioned approvals, data sharing will be made in a secure setting, on a per-case-specific manner, as defined by the chief information security officer of Clalit Health Services. Please submit such requests to RB.

## Contributions

ND, NB and RB conceived and designed the study. ND, NB and DR participated in data extraction and analysis. ND, NB and RB wrote the manuscript. All authors critically reviewed the manuscript. IG, SS and RB supervised the entire study process.

## Supplementary Information

**Supplemental Table 1:**
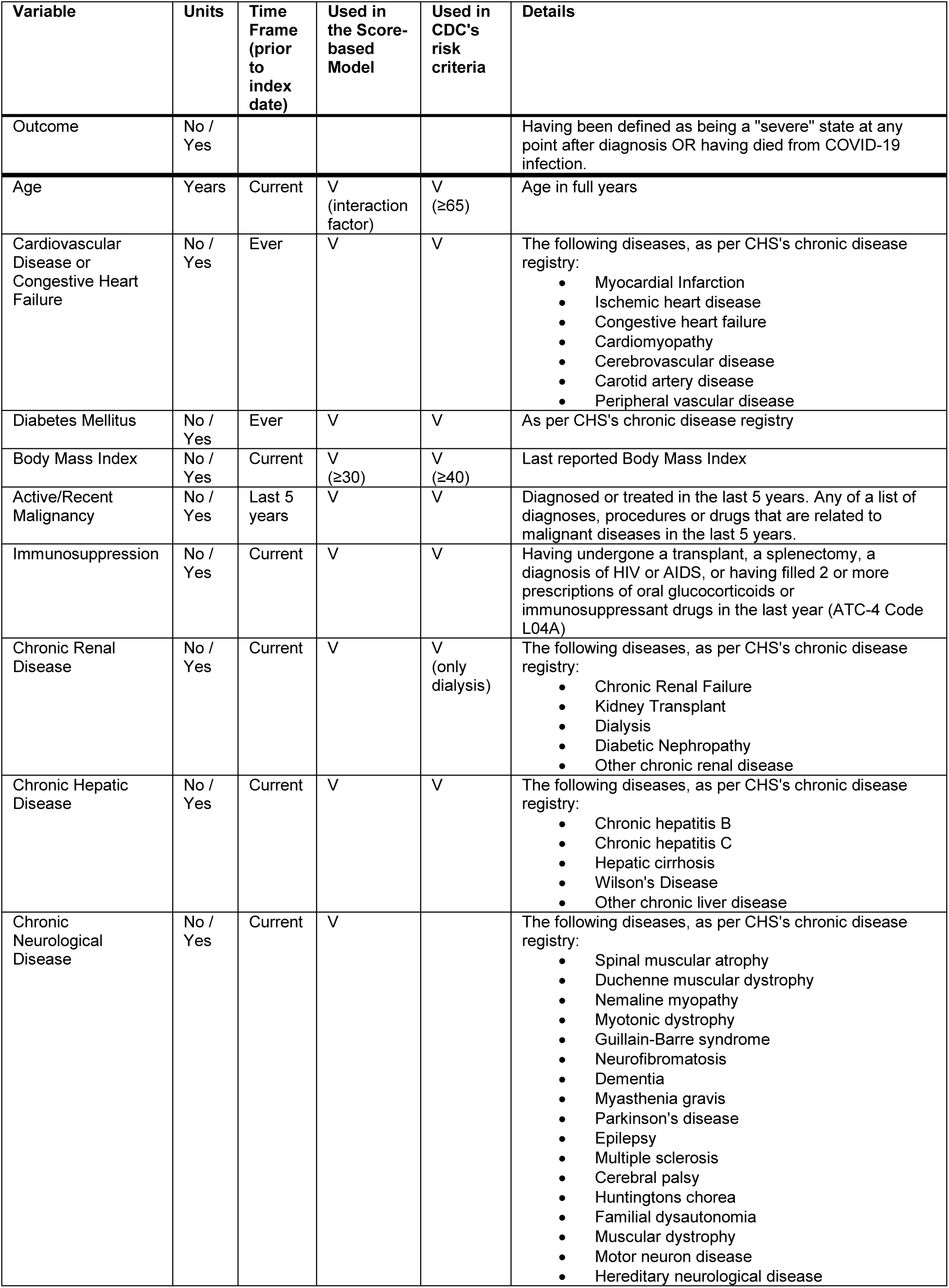
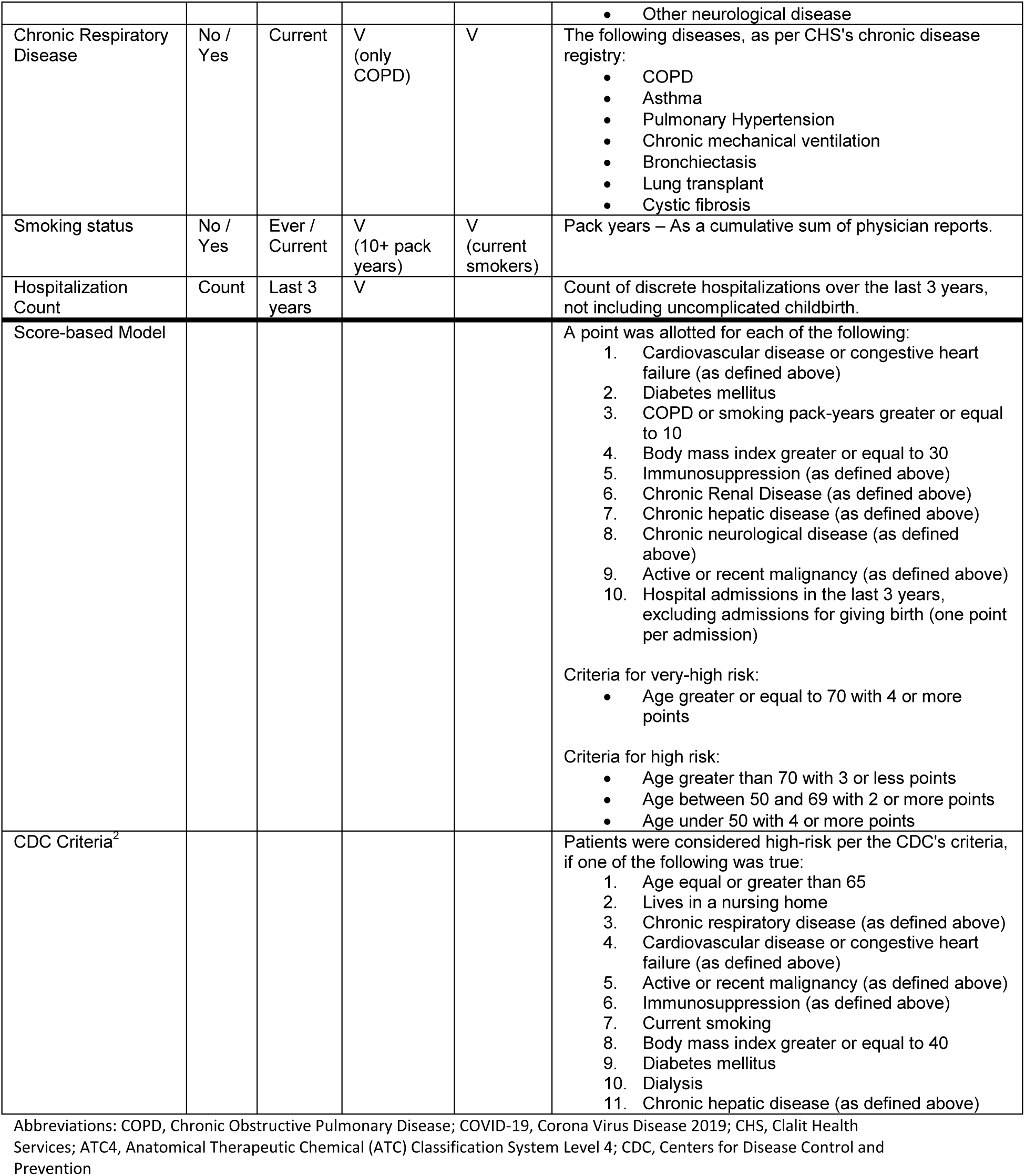
Variable, outcome and model definitions

| Variable | Units | Time Frame (prior to index date) | Used in the Score-based Model | Used in CDC's risk criteria | Details |
| --- | --- | --- | --- | --- | --- |
| Outcome | No / Yes |  |  |  | Having been defined as being a "severe" state at any point after diagnosis OR having died from COVID-19 infection. |
| Age | Years | Current | V (interaction factor) | V ($\geq 65$) | Age in full years |
| Cardiovascular Disease or Congestive Heart Failure | No / Yes | Ever | V | V | The following diseases, as per CHS's chronic disease registry:<br>• Myocardial Infarction<br>• Ischemic heart disease<br>• Congestive heart failure<br>• Cardiomyopathy<br>• Cerebrovascular disease<br>• Carotid artery disease<br>• Peripheral vascular disease |
| Diabetes Mellitus | No / Yes | Ever | V | V | As per CHS's chronic disease registry |
| Body Mass Index | No / Yes | Current | V ($\geq 30$) | V ($\geq 40$) | Last reported Body Mass Index |
| Active/Recent Malignancy | No / Yes | Last 5 years | V | V | Diagnosed or treated in the last 5 years. Any of a list of diagnoses, procedures or drugs that are related to malignant diseases in the last 5 years. |
| Immunosuppression | No / Yes | Current | V | V | Having undergone a transplant, a splenectomy, a diagnosis of HIV or AIDS, or having filled 2 or more prescriptions of oral glucocorticoids or immunosuppressant drugs in the last year (ATC-4 Code L04A) |
| Chronic Renal Disease | No / Yes | Current | V | V (only dialysis) | The following diseases, as per CHS's chronic disease registry:<br>• Chronic Renal Failure<br>• Kidney Transplant<br>• Dialysis<br>• Diabetic Nephropathy<br>• Other chronic renal disease |
| Chronic Hepatic Disease | No / Yes | Current | V | V | The following diseases, as per CHS's chronic disease registry:<br>• Chronic hepatitis B<br>• Chronic hepatitis C<br>• Hepatic cirrhosis<br>• Wilson's Disease<br>• Other chronic liver disease |
| Chronic Neurological Disease | No / Yes | Current | V |  | The following diseases, as per CHS's chronic disease registry:<br>• Spinal muscular atrophy<br>• Duchenne muscular dystrophy<br>• Nemaline myopathy<br>• Myotonic dystrophy<br>• Guillain-Barre syndrome<br>• Neurofibromatosis<br>• Dementia<br>• Myasthenia gravis<br>• Parkinson's disease<br>• Epilepsy<br>• Multiple sclerosis<br>• Cerebral palsy<br>• Huntingtons chorea<br>• Familial dysautonomia<br>• Muscular dystrophy<br>• Motor neuron disease<br>• Hereditary neurological disease |
|  |  |  |  |  | • Other neurological disease |
| Chronic Respiratory Disease | No / Yes | Current | V (only COPD) | V | The following diseases, as per CHS's chronic disease registry:<br>• COPD<br>• Asthma<br>• Pulmonary Hypertension<br>• Chronic mechanical ventilation<br>• Bronchiectasis<br>• Lung transplant<br>• Cystic fibrosis |
| Smoking status | No / Yes | Ever / Current | V (10+ pack years) | V (current smokers) | Pack years – As a cumulative sum of physician reports. |
| Hospitalization Count | Count | Last 3 years | V |  | Count of discrete hospitalizations over the last 3 years, not including uncomplicated childbirth. |
| Score-based Model |  |  |  |  | A point was allotted for each of the following:<br>1. Cardiovascular disease or congestive heart failure (as defined above)<br>2. Diabetes mellitus<br>3. COPD or smoking pack-years greater or equal to 10<br>4. Body mass index greater or equal to 30<br>5. Immunosuppression (as defined above)<br>6. Chronic Renal Disease (as defined above)<br>7. Chronic hepatic disease (as defined above)<br>8. Chronic neurological disease (as defined above)<br>9. Active or recent malignancy (as defined above)<br>10. Hospital admissions in the last 3 years, excluding admissions for giving birth (one point per admission)<br>Criteria for very-high risk:<br>• Age greater or equal to 70 with 4 or more points<br>Criteria for high risk:<br>• Age greater than 70 with 3 or less points<br>• Age between 50 and 69 with 2 or more points<br>• Age under 50 with 4 or more points |
| CDC Criteria<sup>2</sup> |  |  |  |  | Patients were considered high-risk per the CDC's criteria, if one of the following was true:<br>1. Age equal or greater than 65<br>2. Lives in a nursing home<br>3. Chronic respiratory disease (as defined above)<br>4. Cardiovascular disease or congestive heart failure (as defined above)<br>5. Active or recent malignancy (as defined above)<br>6. Immunosuppression (as defined above)<br>7. Current smoking<br>8. Body mass index greater or equal to 40<br>9. Diabetes mellitus<br>10. Dialysis<br>11. Chronic hepatic disease (as defined above) |

Abbreviations: COPD, Chronic Obstructive Pulmonary Disease; COVID-19, Corona Virus Disease 2019; CHS, Clalit Health Services; ATC4, Anatomical Therapeutic Chemical (ATC) Classification System Level 4; CDC, Centers for Disease Control and Prevention

